# Structural brain correlates of childhood trauma with replication across two large, independent community-based samples

**DOI:** 10.1101/2022.06.07.22276081

**Authors:** Rebecca A. Madden, Kimberley Atkinson, Xueyi Shen, Claire Green, Robert F. Hillary, Emma Hawkins, Anca-Larisa Sandu, Gordon Waiter, Christopher McNeil, Mathew Harris, Archie Campbell, David Porteous, Jennifer A. Macfarlane, Alison Murray, Douglas Steele, Liana Romaniuk, Stephen M. Lawrie, Andrew M. McIntosh, Heather C. Whalley

## Abstract

**Introduction:** Childhood trauma and adversity are common across societies and have strong associations with physical and psychiatric morbidity throughout the life-course. One mechanism through which childhood trauma may predispose individuals to poor psychiatric outcomes, such as raised risk of lifetime depression, could be via associations with brain structure. This study aimed to elucidate the associations between childhood trauma scores and brain structure across two large, independent community cohorts.

**Methods:** The two samples comprised (i) a subsample of individuals from Generation Scotland with imaging and in-depth phenotyping, including the CTQ-28 (n=1,024); and (ii) individuals from UK Biobank with imaging and a modified summary CTQ measure (n=27,202). This comprised n=28,226 for mega-analysis. Scans were processed using *FreeSurfer* image processing software, providing cortical and subcortical as well as global brain metrics. Regression models were used to determine associations between these metrics and childhood trauma measures. Associations between childhood trauma measures and psychiatric phenotypes were also explored.

**Results:** Childhood trauma measures associated with lifetime risk of depression diagnosis with similar ORs across cohorts (OR 1.06, 1.23 GS and UKB respectively), which also related to earlier onset and more recurrent course within both samples. There was also evidence for associations between childhood trauma measures and a range of brain structures. Replicated findings included reduced global brain volumes, reduced cortical surface area but not thickness, with highest effects at mega-analysis seen in the frontal (β=-0.0385, SE=0.0048, p_(FDR)_=5.43×10^−15^) and parietal lobes (β=-0.0387, SE=0.005, p_(FDR)_=1.56×10^−14^). At a regional level, one subcortical regional volume in particular – the ventral diencephalon (VDc) – displayed significant associations with childhood trauma measures across the two cohorts and at mega-analysis (β=-0.0232, SE=0.0039, p_(FDR)_=2.91×10^−8^). There was also evidence for associations with reduced hippocampus, thalamus, and nucleus accumbens volumes, however these were not as consistent across cohorts.

**Discussion:** There was strong evidence for associations between childhood trauma and reduced global and regional brain volumes across cohorts. In particular, the presence of an association between childhood trauma and the volume of the VDc (which includes the hypothalamic area), with replication, provides further evidence of the importance of neuroendocrine stress response pathways in links between early life stress and clinical outcomes.

## Introduction

Adverse childhood experiences (ACEs) are reported to affect around 50% of the UK population (1, 2), with similar proportions worldwide (3). The classification of ACEs varies, but typically include abuse and neglect, witnessing domestic or neighbourhood violence, family substance misuse, and parental divorce. ACEs have been found to act in a dose-dependent manner to raise the risk of a broad range of adverse outcomes in adulthood, including poor psychiatric outcomes (4). The childhood trauma questionnaire (CTQ) describes a narrower range of early life experiences, focussing specifically on abuse and neglect, mainly in the home environment (5). It has been repeatedly demonstrated that experiences that fall under this definition of ‘childhood trauma’ (CT) have stronger associations with poor psychiatric outcomes than other types of childhood adversity (CA). It has been shown, for instance, that interpersonal trauma (adolescent sexual or physical abuse) is more strongly associated with lifetime SCID diagnoses than traumatic bereavement during adolescence (6); and that physical neglect, emotional neglect, and sexual abuse show stronger associations with symptom severity among adult psychiatric inpatients than other ACEs such as peer bullying (7).

The biological underpinnings of the relationship between CA and psychiatric disorders are likely to be multifaceted; including chronic dysregulation of the hypothalamic-pituitary-adrenal axis (8), disrupted attachment and emotional development (9), and deviations from typical brain development (10). The latter hypothesis posits that the experience of trauma during sensitive periods of neurodevelopment may lead to structural abnormalities which predispose the adult individual to pathological symptoms (11). A great deal of previous work has focused on describing the neurostructural consequences of CA; the literature has, however, proven to be inconsistent where results fail to replicate study-to-study.

The field to date has often taken a hypothesis led approach; while there can be a strong rationale for examining specific regions of interest (ROIs) in relation to CA, this *a priori* approach can also contribute to bias and inconsistencies, whereby meaningful effects in other regions may be missed. The hippocampus, for instance, has received a lot of attention due to evidence for smaller hippocampal volumes in depression (12), alongside findings from preclinical work indicating the hippocampus is particularly vulnerable to early stressful life events (13). These region-of-interest (ROI) studies generally find reduced hippocampal grey matter (GM) volume (14-20) in CA-exposed groups; however other studies report no association (21-24). A meta-analysis including 17 of these ROI studies looking specifically at the hippocampus did find evidence, however, for reduced volumes of the left, right, and total hippocampus (hedges g=-0.642, -0.616, -0.517 respectively), despite significant between-study heterogeneity, and publication bias in the latter (Egger’s t(df=33)=3.44, p=0.001)(25).

The amygdala has been another focus of ROI analyses of the neurostructural effects of CA, given its role in emotion and fear processing (19, 24, 26). Two meta-analyses of amygdala-specific ROI analyses are however contradictory, one reporting reductions in amygdala volume from meta-analysis of 19 studies (25); the other finding no effect in a meta-analysis of 15 studies (16). Other commonly used ROIs have included the caudate nucleus (22, 23, 27), nucleus accumbens (21, 22), orbitofrontal cortex (18, 22), and anterior cingulate cortex (18, 22-24), with mixed findings.

Despite multiple strands of evidence that traumatic events can affect the structure and function of the thalamus, this region is rarely included as an ROI in *a priori* studies of CA. After traumatic events in adulthood, structural and functional changes have been observed in the thalamus (28-30). Thalamic structural deviations after CA have also been observed (31, 32), and childhood trauma has been associated with functional hyperconnectivity of the thalamus in a transdiagnostic sample (33). In addition to these structural and functional imaging findings, endocrine work has repeatedly shown evidence of dysregulation in the hypothalamic-pituitary-adrenal (HPA) axis in CT-exposed adult populations (34-37); implicating associated thalamic structures as well. The lack of inclusion of the thalamus and hypothalamus in *a priori* structural imaging studies of CA demonstrates the potential to miss interesting regions by taking an ROI-based approach.

While brain-wide (*a priori*) analyses have been conducted, they are rarer and often lack robust sample sizes. One such study revealed an association between CTQ scores and right insular surface area, (38). Meta-analyses of voxel-based morphometry (VBM) approaches to brain-wide studies of CA report reduced GM volumes in the dorsolateral prefrontal cortex, right hippocampus, and right postcentral gyrus (across 19 studies with 1095 adult subjects (25)); and GM volume reductions in temporal, frontal, and midbrain regions, and the left postcentral gyrus (across 12 studies with a total of 693 children, adolescents, and adults (39)). The ENIGMA consortium (40) has recently produced larger-sample, *a priori* analyses of the impact of CA on adult brain structure based on meta-analysed data from multiple cohort studies. In an analysis across 7 subcortical grey matter structures defined by the FreeSurfer image processing software, no significant main effects of childhood adversity were found (41). A similar approach taken in cortical thickness and surface area measures found main effects of CA score in the thickness of the banks of the superior temporal sulcus and the supramarginal gyrus, and the surface area of the middle temporal gyrus (42). They also found various interaction effects with age, sex, and MDD diagnosis.

It is evident that there is limited consistency in determining the relationships between childhood trauma and adult structural imaging findings from whole-brain analyses. These inconsistencies may result from between-study variability. In particular, differences in the detail and definition of childhood trauma/adversity; in the demographics; and in the use of healthy or psychiatric populations (with varying diagnoses and severities of clinical status). With the growing opportunities provided by neuro-imaged population cohorts, large sample sizes are available within a single study protocol, enabling the analysis of neurostructural sequalae of childhood trauma more homogenous samples. In this study, therefore, we describe psychiatric phenotyping, whole-brain analysis, and regional analysis of childhood trauma (CT) in a subsample of the Generation Scotland: Scottish Family Health Study (GS) cohort with neuroimaging data. We based our initial analyses in this cohort due to its characteristically deep phenotyping, including the 28-item Childhood Trauma Questionnaire. We then sought to replicate in the larger UK Biobank cohort, which utilised a much shorter questionnaire assessing CT. In addition, we report imaging mega-analysis from the combined cohorts, with a total sample size of n=28,226. These large samples make the findings presented here well-powered, and more generalisable to the population than smaller clinically oriented samples.

## Methods

The analysis comprised two independent populations: a subsample from The Generation Scotland (n=1,024) and the UK Biobank cohort (n=27,202), totalling a population of n=28,226. In both GS and UKB, freesurfer derived measures of cortical metrics and subcortical volumes were analysed. The analysis focussed initially on global cortical measures, lobar regions, and finally individual regional measures.

### The Generation Scotland cohort

Generation Scotland: Scottish Family Health Study (GS hereafter) is a population-based cohort of over 24,000 individuals with in-depth phenotyping recruited between 2006-2011. A subsample of participants was re-contacted in 2015-2019 for neuroimaging and further data collection. This subsample was used as the basis for the current study, and is described in detail elsewhere (43-46). Cognitive assessments, blood sampling, physical measurements, and clinical questionnaires were also collected from GS participants, including the 28-item Childhood Trauma Questionnaire (CTQ; (5)). The subsample included n=1,153 participants with CTQ and depression phenotyping, and n=1,024 also had MRI imaging data.

The 28-item CTQ is a questionnaire validated for self-report of abuse and neglect during childhood (5). The questionnaire is made up of five subscales – Emotional Abuse (EA), Emotional Neglect (EN), Physical Abuse (PA), Physical Neglect (PN), and Sexual Abuse (SA) – with five items for each subscale, scored on a five-point Likert scale rating frequency of each experience (1-5). Emotional and Physical Neglect items are reverse-scored. The minimisation and denial scale makes up the remaining three items, a scale devised to help detect under-reporting of childhood trauma (5). The analyses reported here focused mainly on the summed score of the 25 subscale items of this questionnaire, scored on a scale 25-125, which provides an index of the cumulative traumatic experience of an individual, consistent with previous literature (e.g. (47, 48)). The questionnaire can also be used to break scores down into severity categories of none-low, low-moderate, moderate-severe, and severe-extreme, for both total CTQ score and each subscale (49, 50).

All analyses were performed using scaled scores for the five childhood trauma subscales captured by the CTQ (and, in UKB, CTM) measures – emotional neglect, emotional abuse, physical neglect, physical abuse, and sexual abuse – as well as scaled composite ‘abuse’ and ‘neglect’ scores created by summing scores on the abuse and neglect items, respectively.

Major Depressive Disorder (MDD) diagnoses were derived from the Structured Clinical Interview for DSM-IV (SCID; (43, 51)) taken at GS baseline recruitment; a binary variable describing presence/absence of a lifetime diagnosis encompasses single-episode, chronic, post-partum onset, and depression with manic/hypomanic features. Two individuals with manic/hypomanic episodes alone were excluded from further analyses. The Quick Inventory of Depressive Symptomatology (52) was used as measure of continuous (and current) depressive symptom severity at the time of the imaging clinic visit.

### MRI Imaging in GS

Imaging was conducted at two sites, Aberdeen and Dundee. In Aberdeen, brain magnetic resonance imaging (MRI) data were acquired on a 3T Philips Achieva TX-series MRI system (Philips Healthcare, Best, Netherlands) with a 32-channel phased-array head coil with a back facing mirror (software version 5.1.7; gradients with maximum amplitude 80 mT/m and maximum slew rate 100 T/m/s). In Dundee, images were acquired on a Siemens 3T Prisma-FIT (Siemens Healthineers, Erlangen, Germany) with 20 channel head and neck coil and a back facing mirror (software version VE11, gradient with max amplitude 80 mT/m and maximum slew rate 200 T/m/s). The current study uses T1 structural data, though other sequences were also acquired (53, 54). Structural measures were derived in-house from raw images using FreeSurfer version 5.3 (55-57). The 1,070 images were segmented into grey matter, white matter, and cerebrospinal fluid. Grey matter was further segmented into cortical and subcortical grey matter. The cortex was divided into 34 regions per hemisphere according to the Desikan-Killany atlas (58). Visual quality control was undertaken for exclusion of participants with major output errors in segmentation or parcellation. There were 424 subjects for whom a degree of manual editing was required to make small corrections to the images. Participants (n=12) were excluded if errors could not be corrected in this way. For the 68 bilateral cortical regions, measures of mean thickness, surface area, and volume were calculated. For 8 bilateral subcortical grey matter structures, volume alone was calculated. Coordinates of head position within the scanner were included as covariates.

### The UK Biobank cohort

UK Biobank (UKB) is a large, UK-based population cohort of over n=500,000 adults recruited between 2006-2010 (59). Data was collected over several instances of postal questionnaires and clinic visits. MRI imaging was performed on n=100,000, which was acquired in-clinic during the second assessment instance (59, 60); at the time of writing, MRI data were available for n=41,985 (61). An online follow-up questionnaire was disseminated to participants after completion of the imaging appointment, comprising the Mental Health Questionnaire (MHQ; which included childhood trauma questions).

Five items relating to childhood trauma were included in the MHQ, derived from the 28-item CTQ with each item scored on a five-point likert scale (0-4). Hence only one item related to each of the five subscales (as described in the fuller 28-item CTQ) was included in this cohort, making this data less sensitive than the GS and necessitating a focus throughout the study on total scores (with subscale results presented in Appendix 3). For the purposes of this study, childhood trauma items were used on a 0-20 scale of the summed total of responses to these items. Any individuals with incomplete responses to the childhood trauma items were excluded from analyses, leaving a maximum sample of n=153,650 for this report, of which n=27,202 had MRI imaging data available.

MDD diagnoses were derived from ICD10 diagnostic criteria assessed at the initial assessment centre visit (62), and CIDI-SF (63) derived questionnaire in the MHQ at online follow up. A modified form of the Generalised Anxiety Disorder Assessment (GAD-7; (64)) was delivered in the MHQ, as well, to indicate recent anxiety symptoms. The 4-item Patient Health Questionnaire (PHQ-4 (62)) was used to take current mood level at the assessment centre appointment.

### MRI Scanning in UKB

Imaging for the 40,000 data release in January 2020 was conducted at three sites: Stockport, Newcastle, and Reading. These sites are equipped with identical MRI scanners - 3T Siemens Skyra (software VD13), using the standard Siemens 32-channel head coil (65). The current study uses T1 structural scans, although other sequences were also collected. Structural measures were derived for the 40,000 data release using FreeSurfer software (61). See supplementary information p.3

### Statistical analyses

All statistical analyses were performed in R v.3.6.4. All analyses were corrected for multiple comparisons using the false-discovery rate (FDR) method.

Linear and binomial regressions were used to describe phenotypic relationships between scaled childhood trauma and demographic information, as well as features of depressive symptomatology in both cohorts. Numbers for each model varied in both cohorts, as different measures had different numbers of non-completers. These models adjusted for age and sex as minimum covariates.

To determine the relationships between scaled childhood trauma total and subscale scores and structural brain imaging features, linear regression models were conducted using the NLME package in R. For cortical structures, volume, surface area, and thickness measures were available; for subcortical structures volume data were available. Covariates included sex, age, age squared (to account for non-linear effects of age), scaled intracranial volume, hemisphere, scan site. For 5 summed lobar measures (lobar cortical volume, lobar cortical surface area, and mean lobar cortical thickness), the same covariates were used. For global measures (such as whole brain volume (WBV), total grey matter volume, and total white matter volume), hemisphere was not a required covariate and ICV was also excluded at a covariate due to high collinearity with the measures.

Imaging analyses were performed in n=1,024 in GS, and n=27,202 (n=26,639 for WBV) in UKB. These numbers represent those with complete imaging data as well as complete childhood trauma data.

### Mega-analysis of imaging samples

For the mega-analysis, the total and subscale childhood trauma scores were scaled within each cohort, and then re-scaled across the merged mega-analysis sample. Structural imaging data including ICV was scaled within cohort. Analyses were conducted, as before, using the same covariates.

## Results

### Demographics characteristics

#### Generation Scotland

The GS study population (n=1,153) was 58.54% female, with a mean age of 59.52yrs (26-84yrs). At SCID interview 30.79% met criteria for lifetime depression (Table 1).

**Table 1:** Demographic characteristics and psychiatric traits of the GS study population, incorporating participants with complete CTQ responses and SCID interviews. Prevalence of the 5 CTQ subscales are also shown. *\* For the n=355 reporting lifetime depression*

| Trait | total N | Prevalence |  |
| --- | --- | --- | --- |
|  |  | % |  |
| Sex (female) | 1,153 | 58.54 |  |
| Lifetime MDD | 1,153 | 30.79 |  |
| Recurrent MDD ($\geq 2$ episodes)* | 355 | 62.54 | |
| Any CTQ Subscale > 'low' | 1,153 | 51.00 |  |
| Total CTQ Score > 'low' | 1,153 | 26.02 |  |
| Ever Smoker | 938 | 43.71 |  |
|  |  | mean | range |
| Age | 1,153 | 59.52 | 26-84 |
| BMI (kg/m<sup>2</sup>) | 1,153 | 28.21 | 16.42-64.24 |
| Age of Depression Onset (yrs)* | 355 | 33.45 | 5-65 |
| Subscales |  | Prevalence |  |
|  |  | % |  |
| Emotional Abuse Score > 'low' | 1,153 | 21.25 |  |
| Emotional Neglect Score > 'low' | 1,153 | 31.92 |  |
| Physical Abuse Score > 'low' | 1,153 | 13.54 |  |
| Physical Neglect Score > 'low' | 1,153 | 21.77 |  |
| Sexual Abuse Score > 'low' | 1,153 | 13.80 |  |

Using the CTQ score cut-offs (see supplementary materials, page 1), 26.02% of the study population reported experiencing ‘low-moderate’ levels of childhood trauma, or above. Using the subscale-specific cut-offs, 51% of the study population reported a score of ‘low-moderate’ or above on at least one CTQ subscale. The most frequent subtype of trauma reported was emotional neglect, and the least frequent were physical and sexual abuse (Table 1).

Total CTQ scores were significantly higher in females (mean=34.75) than males (mean=32.94; t_(df=1,138.8)_=2.79, p=0.0054), and were associated with higher BMI (β=0.97, SE=0.17, p_(FDR)_=2.31×10^−8^), but were not significantly associated with age (β=-4.04×10^−4^, SE=0.0029, p_(FDR)_ =0.89).

Participants with lifetime MDD diagnoses reported higher total CTQ scores (mean=39.59) than healthy controls (mean=31.52; t_(df=442.54)_=-9.11, p<0.001). A significant positive association was found between total CTQ score and risk of lifetime depression (β=0.68, OR=1.98, p_(FDR)_=6.45×10^−18^). Depression risk was positively associated with total scores on all trauma subscales (Appendix 1). Higher total CTQ scores also predicted younger age-of-onset for depression (β=-3.2, 95% CI=-4.22 – - 2.17, p_(FDR)_=1.44×10^−8^), and higher odds for a recurrent course of depressive illness (β=0.36, OR=1.43, p_(FDR)_=0.0022).

#### UK BioBank

The UKB study population (n=153,650) was 56.31% female, with a mean age of 55.91 years (38-72yrs). Of n=94,379 participants for whom ICD-10 coded diagnostic information was available, 2.68% report lifetime depression. For the n=123,355 participants who completed a CIDI diagnostic questionnaire, however, 29.47% met criteria for lifetime depression (Table 2).

**Table 2:** Demographic characteristics and psychiatric traits of the UKB study population, incorporating participants with complete childhood trauma responses. *\* Data comes from initial assessment centre visit, not tied to a diagnostic questionnaire*. *\* * Data comes from online follow-up to imaging appointment, not tied to a diagnostic questionnaire*.

| *Trait* | **total N** | **Prevalence** |  |
| --- | --- | --- | --- |
|  |  | **%** |  |
| Sex (female) | 153,650 | 56.31 |  |
| Lifetime MDD (ICD 10) | 94,379 | 2.68 |  |
| Lifetime MDD (CIDI) | 123,355 | 29.47 |  |
| Recurrent MDD (≥2 episodes)* | 23,004 | 69.4 |  |
| Any CTQ Subscale > 'Rarely True' | 153,650 | 59.28 |  |
| Ever Smoker | 153,306 | 59.68 |  |
|  |  | **mean** | **range** |
| Age | 153,650 | 55.91 | 38-72 |
| Total CTM score | 153,650 | 1.76 | 0-20 |
| BMI (kg/m2) | 153,269 | 26.77 | 12.12-67.38 |
| Age of Depression Onset (yrs)** | 79,288 | 37.24 | 2-78 |
| *Subscales* |  | **Prevalence** |  |
|  |  | **%** |  |
| Emotional Abuse item < 'V. Often True' | 153,650 | 14.41 |  |
| Emotional Neglect item > 'Rarely True' | 153,650 | 47.59 |  |
| Physical Abuse item > 'Rarely True' | 153,650 | 16.22 |  |
| Physical Neglect item < 'V. Often True' | 153,650 | 16.21 |  |
| Sexual Abuse item > 'Rarely True' | 153,650 | 8.76 |  |

Due to the less detailed nature of the childhood trauma metric (CTM) items in UKB’s MHQ, the severity cut-offs inherent to the CTQ could not be replicated. The proportion of participants responding higher than ‘Rarely True’ (or lower than ‘Very Often True’ for reverse-coded items) to any CTM item was 59.28% (Table 2).

Total CTM scores in UKB were significantly higher in females (mean=1.85) than in males (mean=1.64; t_(df=152,796)_=16.85, p<0.001). They showed a positive association with BMI (β=0.37, SE=0.012, p_(FDR)_=1.86×10^−220^), and a small but significant negative association with age (β=-0.0059, SE=3.3×10^−4^, p_(FDR)_=3.28×10^−72^).

Participants with CIDI-coded lifetime MDD had higher CTM scores (mean=2.57) than healthy controls (mean=1.32; t_(df=48,247)_=-72.35, p<0.001); similarly, those with ICD-10 coded lifetime MDD had higher CTM scores (mean=3.34) than healthy controls (mean=1.67; t_(df=2582)_=-22.18, p<0.001). Significant positive associations were found between total CTM scores and risk of lifetime depression for CIDI (β=0.49, OR=1.63, p_(FDR)_<1×10^−314^), and ICD-10 data (β=0.4, OR=1.49, p_(FDR)_=8.52×10^−186^). ICD-10- and CIDI-diagnosed depression risk both positively associated with responses to each of the CTM subscale items, in addition (Appendix 1). Total CTM scores predicted younger age of depression onset (β=-2.1, SE=0.046, p_(FDR)_<1×10^−314^), and higher odds of a recurrent course of depressive illness (β=0.22, OR=1.24, p_(FDR)_=2.66×10^−51^).

### Neuroimaging findings

We took the approach of analysing relationships between scaled CT scores and brain metrics in increasing levels of detail, starting with global brain and grey/white matter volumes, then lobar-level metrics, then across all regions.

#### Global brain measures

The relationships between CT scores and global brain metrics were analysed in GS (n=1,024), UKB (n=26,639), and the mega-analysis sample (n=28,226).

In GS, significant negative associations were revealed between total CTQ score and global white matter volume (β=-0.0852, SE=0.0258, p_(FDR)_=0.0015), global grey matter volume (β=-0.0682, SE=0.0219, p_(FDR)_=0.0019), and whole brain volume (β=-0.0844, SE=0.0239, p_(FDR)_=0.0013; Appendix 2).

In UKB, CTM scores showed significant negative associations with global white matter volume (β=-0.0311, SE=0.0049, p_(FDR)_=2.67×10^−10^), global grey matter volume (β=-0.0433, SE=0.0049, p_(FDR)_=5.27×10^−18^), and whole brain volume (β=-0.0399, SE=0.0048, p_(FDR)_=1.75×10^−16^; Appendix 2).

In the mega-analysis population, the whole-brain analyses followed much the same pattern, with significant negative associations between childhood trauma scores and global white matter volume (β=-0.0330, SE=0.0048, p_(FDR)_=9.15×10^−12^), global grey matter volume (β=-0.0442, SE=0.0048, p_(FDR)_=1.74×10^−19^), and whole brain volume (β=-0.0417, SE=0.0047, p_(FDR)_=1.51×10^−18^; Appendix 2).

#### Lobar measures

The relationships between scaled CT scores and the whole-lobe cortical volume, whole-lobe cortical surface area, and mean cortical thickness of all five lobar regions were examined and analysed in GS (n=1,024), UKB (n=27,202), and the mega-analysis sample (n=28,226).

In GS, there were significant negative associations between total CTQ scores and both cortical surface areas and cortical volumes of the frontal, parietal, temporal, occipital, and cingulate lobes (Appendix 2). There were no significant associations for lobar cortical thicknesses.

In UKB, we observed significant negative associations between total CTM score and cortical volumes and cortical surface areas of all five lobes, but no associations with cortical thicknesses were observed (Appendix 2).

In the mega-analysis sample, relationships between childhood trauma score and cortical volume and surface areas of all five lobes were significant (Appendix 2). Again, no associations between childhood trauma scores and whole-lobe cortical thickness were observed.

#### Regional measures

Regional analyses examined the relationships between CT scores and volumes of 34 cortical and 8 subcortical regions, as well as thickness and surface area for the same 34 cortical regions in GS (n=1,024). In UKB (n=27,202) and mega-analysis (n=28,226), 33 cortical regions were examined on the same metrics, plus 8 subcortical regional volumes.

In GS, across all regional metrics analysed three regions were significantly associated after FDR correction (**Figure 2**, Appendix 2). These were the volumes of the hippocampus (β=-0.0582, SE=0.0247, p_(FDR)_=0.050), nucleus accumbens (β=-0.0654, SE=0.0231, p_(FDR)_=0.037), and ventral diencephalon (VDc; **Supplementary Figure 1**; β=-0.0526, SE=0.0218, p_(FDR)_=0.050). All three regions are subcortical areas, and all were negatively associated with total CTQ score, with higher CTQ score predicting lower volume of these regions.

**Figure 1:**
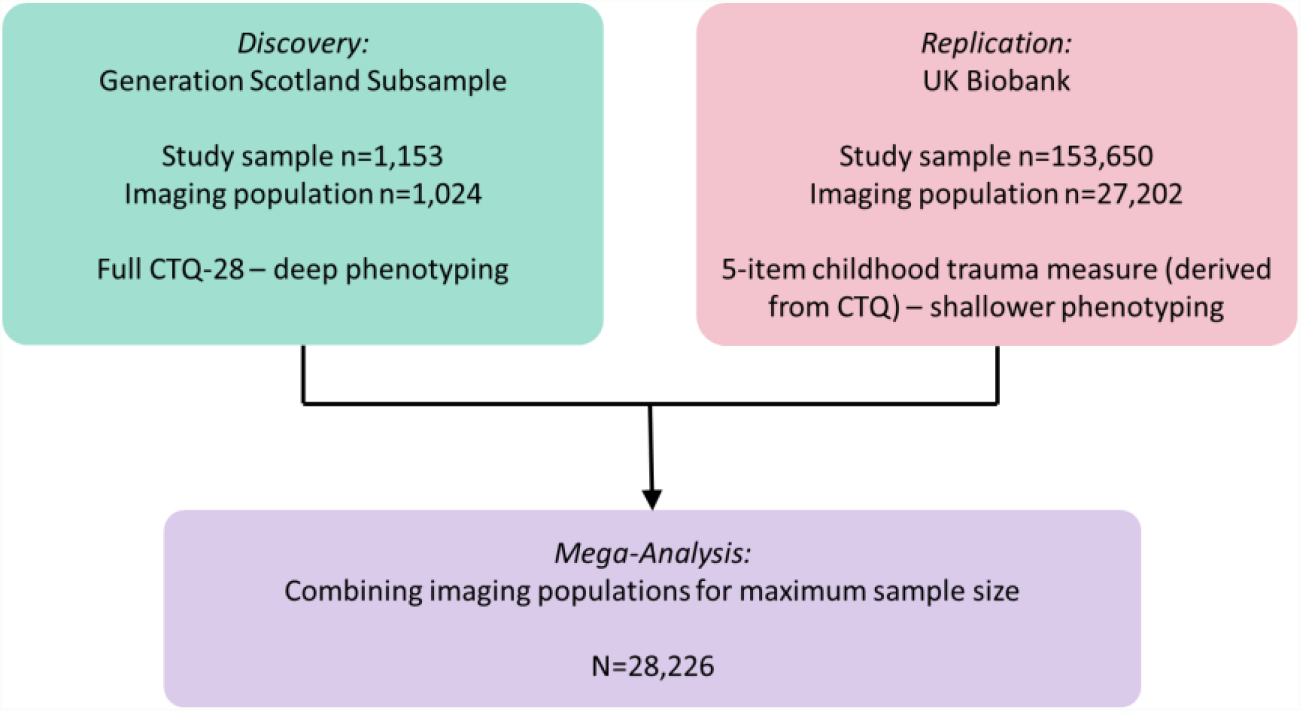
Analysis structure of the present study, demonstrating the initial discovery analysis in GS due to deeper phenotyping of childhood trauma, with replication in UKB and mega-analysis for maximum sample size.

**Figure 2:**
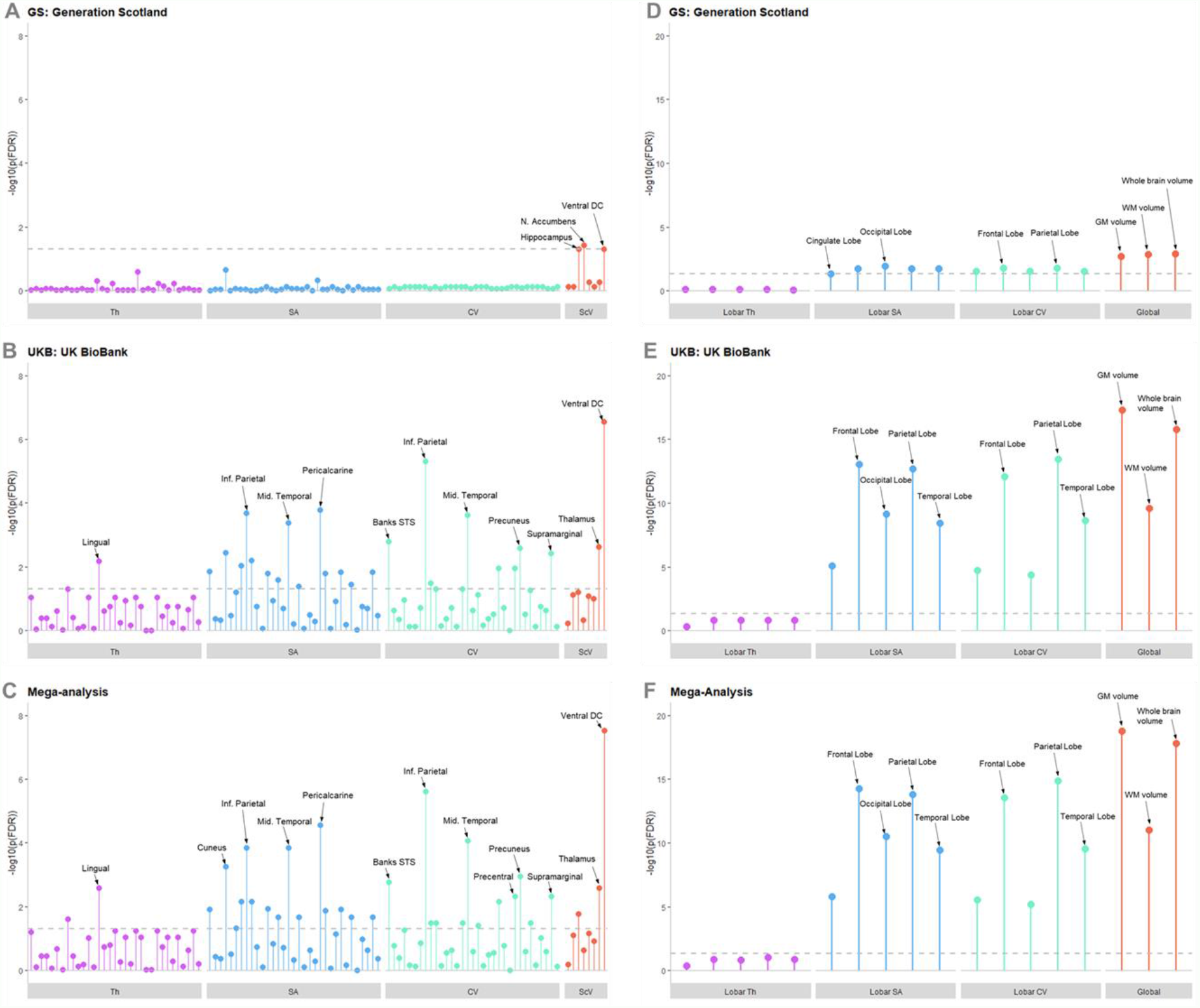
Lollipop plots showing –log10 of p_(FDR)_ for each region and metric in the regional analysis for (A) GS, (B) UKB, and (C) the mega-analysis; and in the global and lobar analyses for (D) GS, (E) UKB, and (F) the meg-analysis. The –log10 of p_(FDR)_=0.05 is represented by the dotted grey line, any points exceeding this line achieved statistical significance after FDR correction. Regions of higher significance are labelled, the order in which other regions are presented can be found in Appendix 2. *In this figure only: Th = Cortical thickness, SA = Cortical surface area, CV = Cortical volume, ScV = Subcortical Volume*.

In UKB, many significant associations were found (**Figure 2;** Appendix 2), due to the higher power afforded by the larger sample size in this data set. Of note was the replication of a significant negative association between total CTM score and the volume of the VDc (β=-0.0220, SE=0.0040, p_(FDR)_=2.84×10^−7^). Significant associations between CTM scores and cortical regions were demonstrated – again showing more consistent associations with SA and volume than cortical thickness. Regions with the strongest effect sizes were the volume of the inferior parietal lobule (β=-0.0223, SE=0.0042, p_(FDR)_=4.78×10^−6^), and the surface area of the pericalcarine cortex (β=-0.0248, SE=0.0054, p_(FDR)_=1.61×10^−4^).

All findings from the UKB analyses were replicated in the mega-analysis, which also revealed some novel significant associations (**Figure 2, Figure 3**, Appendix 2). Two of the three findings in the GS regional analysis were also replicated in the mega-analysis; the significant negative associations in the hippocampus (β=-0.0119, SE=0.0044, p_(FDR)_=0.017), and the ventral diencephalon (β=-0.0232, SE=0.0039, p_(FDR)_=2.91×10^−8^). The volume of the thalamus was additionally significant at mega-analysis (β=-0.0134, SE=0.0039, p_(FDR)_=0.0026).

**Figure 3:**
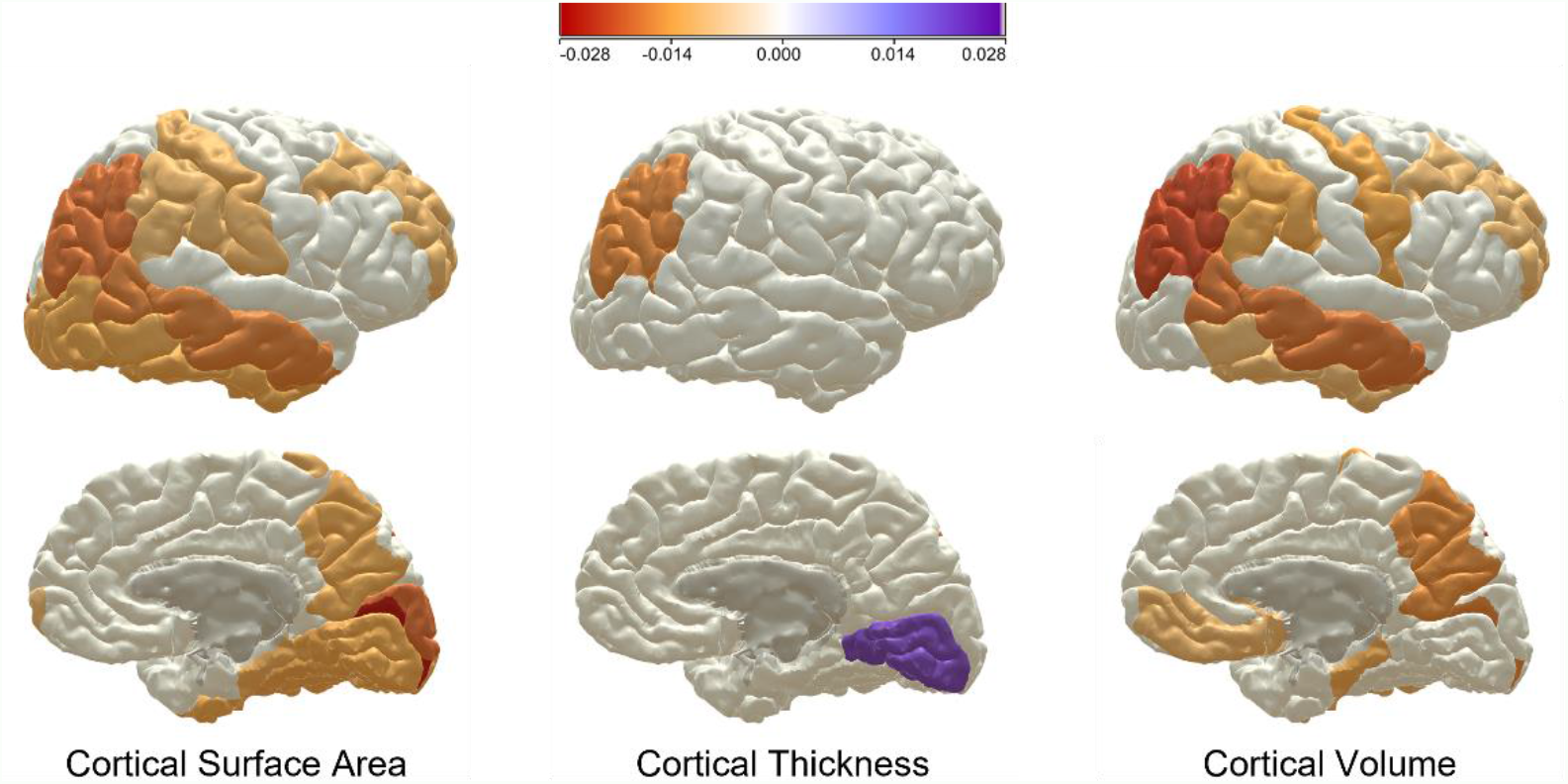
Map of showing Beta values for regions significantly associated with childhood trauma in the mega-analysis, for cortical thickness, cortical surface area, and cortical volume. The red-toned colours represent a negative association and the purple-toned represent a positive association, with the strength of that associated denoted by the shade of the colour. Images created using the Cox & Liewald *Heatmapper* programme (66)

#### Childhood trauma subscales

Across all three global brain metrics in GS, significant associations were demonstrated with Emotional Abuse, Physical Abuse, Sexual Abuse, and Abuse composite score. This was replicated in the UKB and mega-analysis. In GS, however, no significant association was indicated for Emotional Neglect, Physical Neglect, or Neglect Composite score. This was not replicated in the UKB and mega-analysis; EN, PN, and Neglect composite were significantly associated with all three global brain metrics in these analyses (Appendix 3).

Similarly, at the lobar level, imaging samples demonstrated associations with some subscale scores with particularly consistent results between PA, SA, and abuse composite score and lobar surface areas; and between PA and abuse composite score and lobar volumes. Very few significant associations were found with lobar thicknesses, excepting significant associations with both abuse and neglect composite scores in the UKB analysis only (Appendix 3). At the regional level in GS, the only significant associations observed were between the physical abuse score and the surface area of superior parietal cortex (β=-0.0771, SE=0.0235, p_(FDR)_=0.036), the cortical volume of the entorhinal cortex (β=-0.0733, SE=0.0224, p_(FDR)_=0.037), and 6 of the 8 subcortical volumes: the hippocampus, VDc, thalamus, amygdala, nucleus accumbens, and pallidum (Appendix 3). No other subscale or composite score showed any significant associations with any regional metric in GS, except for the neglect composite score which associated significantly with the volume of the nucleus accumbens (β=-0.0805, SE=0.0229, p_(FDR)_=0.0037).

In UKB and in the mega-analysis, many associations were found between the different trauma subtypes and regional imaging metrics, including some associations between the emotional abuse items and cortical thickness measures (Appendix 3). The abuse composite score demonstrated significant associations with the volumes of the hippocampus, thalamus, and ventral diencephalon in both the UKB analysis and mega-analysis – the same three subcortical regions with which total childhood trauma scores associated in the mega-analysis (Appendix 3). The only region for which a significant association was found across all three analyses was the VDc (GS: β=-0.0676, SE=0.0217, p_(FDR)_=0.005; UKB: β=-0.0212, SE=0.004, p_(FDR)_=1.01×10^−6^; Mega-analysis: β=-0.023, SE=0.0039, p_(FDR)_=4.19×10^−8^).

## Discussion

This study reports psychiatric and structural brain correlates of childhood trauma in a mega-analysis of two independent UK cohorts. The mega-analysis sample size of n=28,226 improves significantly upon the sample sizes of similar multicentre studies (41, 42).

The data presented here provide further validation for proposed relationships between childhood trauma and depressive pathology in adulthood. These links to poor mental health outcomes have been demonstrated previously (67), as have links to poor physical health outcomes (4). The study also highlights the high prevalence of traumatic childhood experiences in our population, with 51% and 59% of participants reporting having experienced some kind of adverse childhood experience in GS and UKB, respectively. In the GS CTQ data, where more detailed analysis was possible, we observed what might be considered a ‘clinically significant’ level of trauma (a total CTQ score over the ‘low-moderate’ threshold) in 26% of the study population. This fits with previous estimates of childhood trauma prevalence (1, 2, 4). The association between higher CT score and younger age of onset for MDD is strong in both cohorts (GS: β=-3.2, 95% CI=-4.22 – -2.17, p_(FDR)_=1.44×10^−8^; UKB: β=-2.1, SE=0.046, p_(FDR)_<1×10^−314^). This replicated indicates that CT is having early effects on the development of psychopathology, and given the links between age of onset and poorer long-term outcomes in MDD could also suggest a link with lifetime severity (68).

The severity of traumatic childhood experiences associated with global brain volume, global grey and white matter volumes, and the volumes and surface areas of all five major brain lobes at mega-analysis with effect sizes ranging from -0.0442 – -0.0204 (Appendix 2). These results point to a whole-brain effect of adverse childhood events on brain development, perhaps with exacerbated results in particular regions. Interestingly, in the global, lobar, and regional analyses very few associations are seen between CT scores and cortical thickness metrics. The evidence of this work suggests that CT severity may be more strongly associated with cortical surface areas and volumes, especially in front and temporo-parietal regions (**Figure 3**). Specific reductions in cortical surface area rather than thickness have also been reported in the context of adolescent depression (69), this could reflect a common feature associated with early onset of psychiatric morbidity. Cortical surface area has been shown to be established earlier in development within a shorter window than cortical thickness (70); adverse experiences within this earlier window could be interrupting the developmental process resulting in the surface area findings presented here. Cortical thickness is established over a much longer time period, well into adulthood (70), and may therefore be less susceptible to interruption by early life events.

The effect sizes found in the global and lobar analyses presented here are larger than those found in the regional analyses, but there were nevertheless multiple significant associations demonstrated. Total CT scores was significantly associated with multiple cortical and subcortical regional volumes at mega-analysis, with effect sizes ranging from -0.0262 – 0.0208. The strongest effect sizes were seen in the negative relationship between total childhood trauma scores and the volume of the ventral diencephalon (β=-0.0232, SE=0.0039, p_(FDR)_=2.91×10^−8^), the volume of the inferior parietal cortex (β=-0.0225, SE=0.0042, p_(FDR)_=2.43×10^−6^), and the surface area of the pericalcarine cortex (β=-0.0262, SE=0.0053, p_(FDR)_=2.75×10^−5^); as well as the positive association with the thickness of the lingual cortex (β=0.0208, SE=0.0053, p_(FDR)_=0.0026). The subcortical regions the hippocampus (β=-0.0119, SE=0.0044, p_(FDR)_=0.017) and the thalamus (β=-0.0134, SE=0.0039, p_(FDR)_=0.0026) were also significantly associated with total CT scores at mega-analysis, and in the GS analysis the nucleus accumbens was significant with a relatively strong effect size (β=-0.0654, SE=0.023, p_(FDR)_=0.037). We have previously reported an association between elevated hair glucocorticoids and the volume of the nucleus accumbens (as well as with MDD and CT) in the GS imaging sample, suggesting a possible link to the HPA axis and stress reactivity (71).

The ventral diencephalon was the only regional metric in which a consistent effect was seen across both GS and UK Biobank, as well as in the mega-analysis. The VDc is a *FreeSurfer* derived region comprising several structures, including the hypothalamus – a region crucial to the regulation of the stress response – as well as subthalamic nuclei, the substantia nigra, the medial and lateral geniculate nuclei and associated white matter structures (**Figure 2**; as described (72, 73)). A significant relationship was also observed between CT and the volume of the thalamus (β=-0.0134, SE=0.0039, p_(FDR)_=0.0026). This could implicate involvement of thalamic/hypothalamic function after CT, supporting previous work (31-37). Future work could employ finer-grained approaches to probe associations with this region more deeply.

It is well established that different types of trauma may affect the brain in different ways (10). The subscale mega-analysis presented here generally found an influence of physical abuse on a greater number of regional surface areas and cortical volumes than other subscales, although physical neglect also associated with a high number of regional surface areas (Appendix 3). Emotional abuse was the only subscale which had any associations with regional cortical thicknesses (Appendix 3). Subcortical regional volumes were affected by all subscales of childhood trauma, with the abuse composite measure showing the same pattern of effects as the total CT score – negatively associating with the hippocampus (β=-0.0124, SE=0.044, p_(FDR)_=0.013), thalamus (β=-0.0166, SE=0.0040, p_(FDR)_=1.03××10^−4^), and VDc (β=-0.0249, SE=0.0040, p_(FDR)_=2.42×10^−9^) – indicating that abuse may be more strongly associated with structural brain changes after CT than neglect. It is important, however, to treat the UKB and mega-analysis subscales with caution due to the single-factor nature of the subscale items in UKB. In GS alone, due to the lower power of the cohort in comparison, few significant associations were found in the subscale analyses. Notably, physical abuse associated significantly with the surface area of the superior parietal cortex (β=-0.0771, SE=0.024, p_(FDR)_=0.036), and the volume of the entorhinal cortex (β=-0.0733, SE=0.022, p_(FDR)_=0.037), as well as 6 of 8 subcortical regions (Appendix 3). No other subscale associated with any cortical measures in GS, although physical and emotional neglect both associated with the volume of the nucleus accumbens (β=-0.0681, SE=0.023, p_(FDR)_=0.025; β=-0.0743, SE=0.023, p_(FDR)_=0.0099 respectively), for which the neglect composite score was accordingly significant (β=-0.0805, SE=0.023, p_(FDR)_=0.0037). In the global and lobar level analyses in GS, physical and sexual abuse associated with most lobar surface areas, and physical abuse associated with most lobar volumes (Appendix 3). The physical, emotional, and sexual abuse items and the abuse composite score each associated with the global brain volume, and global grey and white matter measures (effect sizes ranging from -0.114 --0480; Appendix 3); meanwhile, neither neglect item not the composite score associated with any global measure in GS alone. This suggests that the effect of childhood trauma on global brain development may be more influenced by experiences of abuse than neglect, with physical abuse seeming to display the strongest influence.

### Limitations

Despite the very strong sample size derived from mega-analysis of two independent UK cohorts, enabling well-powered brain-wide analysis and conservative correction for multiple testing, the multi-sample nature of this study introduces unavoidable limitations. Chiefly the use of different childhood trauma measures, upon which this work relies heavily. The GS subsample employed the full CTQ-28, while the UKB study used a five-item CT metric comprising one item taken from each subscale of the CTQ-28. While this does mean that the two measures share features, they are not precisely equivalent. The cruder nature of the UKB measure is evident in the high report of emotional neglect in UKB (47.59% responding ‘rarely true’ or above, compared to 31.92% reporting a ‘low-moderate’ score or above in GS). The single item used to denote emotional neglect in UKB was ‘When I was growing up… I felt loved’, score ‘never true’ to ‘very often true’. This item carries a high emotional valence, which may have led respondents to over-report compared to the scores derived from a multi-factorial scale in the full CTQ-28. The subscale analyses presented here should be treated particularly cautiously as a result, in both UKB and in the mega-analysis sample due to the heavy representation of UKB participants there. Other metrics used in the two cohorts – such as psychiatric diagnostic tools – differed, leading again to a degree of mismatch in the data derived from each cohort. The imaging protocols, too, were conducted at different sites and scanners and according to different protocols, although the ‘scan site’ variable employed in all imaging analyses attempts to account for this. Despite these limitations, however, the two samples were relatively homogenous compared to previous multi-cohort studies, as can be seen by comparing the study population demographics presented for each cohort (**Table 1, Table 2**).

Further limitations are evident in the nature of the CTQ in itself – retrospective report of traumatic childhood experiences has been shown to identify a different population than prospective report (74). While it is difficult to conceive of a study on this scale that does not rely on retrospective report, it is important to consider these results in light of this fact. Neither cohort provides any information on the timing of reported trauma, either. This has been shown to affect outcomes, with different types of trauma affecting brain development in different ways according to the timing of these events (75). An additional concern relevant to any population imaging cohort is evidence that ‘healthy bias’ may exist in the group of participants who were able and willing to attend MRI scanning appointments as compared to the un-scanned members of the cohort. Healthy bias has been demonstrated in UKB (76) but may well be present in GS, thus participants with more extreme symptoms following CT may not have been included in the study populations reported here.

### Conclusions

This large mega-analysis study reports associations between childhood trauma and altered structural metrics across the adult brain, with analyses performed brain-wide in a non-hypothesis driven manner. These effects can be seen at the global, lobar, and regional levels. A negative association between childhood trauma metric scores and the ventral diencephalon – an area comprising the hypothalamus and other thalamus-associated regions – replicated across all three analyses. The effects on global brain measures are particularly noteworthy, and support evidence of an involvement of traumatic childhood experiences in global brain development.

## Data Availability

All data produced in the present study are available upon reasonable request to the authors

## List of Abbreviations

CA: Childhood adversity
ACEs: Adverse childhood experiences
CT: Childhood trauma
CTQ: Childhood trauma questionnaire
CTM: childhood trauma metric (shorthand for UKB childhood trauma items)
SCID: Structured clinical interview for DSM-IV
VDc: Ventral diencephalon
EN: Emotional Neglect
EA: Emotional Abuse
PN: Physical Neglect
PA: Physical Abuse
SA: Sexual Abuse

