## Supplementary material for "Structural brain correlates of childhood trauma with replication across two large, independent community-based samples": Suplementary Information

**Supplementary Information**

CTQ-28 – further scoring information:

CTQ scores can be split into severity categories, as used in previous work (1, 2).

The cut-off scores vary between subscales, they are:

- None/Minimal (hereafter “none”; total CTQ ≤36; EA <8; EN <9; PA <7; PN <7; SA 5)
- Low to Moderate (hereafter “low”; total CTQ 36-51; EA 9-12; EN 10-14; PA 8-9; PN 8-9; SA 6-7)
- Moderate to Severe (hereafter “moderate”; total CTQ 51-68; EA 13-15; EN 15-17; PA 10-12; PN 10-12; SA 8-12)
- Severe to Extreme (hereafter “extreme”; total CTQ ≥69; EA ≥16; EN ≥18; PA ≥13; PN ≥13; SA ≥13)

The Ventral Diencephalon:

The ventral diencephalon is a region produced by Freesurfer subcortical parcellation. It is defined in the FreeSurfer wiki page (3) which cites several references to the structure to support the parcellation (4-6). The ventral diencephalon is also briefly mentioned in the paper describing the parcellation process upon which FreeSurfer is based (7). A commercial company ‘Neuromorphometrics, Inc.’ which utilises FreeSurfer for private ‘labelling’ of MRI scans provides more detail on the structure contained within the ventral diencephalon (although the origin of their information and the overlap with the FreeSurfer region are not made clear). They state:
*“The ventral diencephalon (VDC) is not an anatomical name for a single structure but a name we've given to a group of structures that generally cannot be distinguished from each other with standard MRI images. This "miscellaneous" area includes the hypothalamus, mammillary body, subthalamic nuclei, substantia nigra, red nucleus, lateral geniculate nucleus (LGN), and medial geniculate nucleus (MGN). White matter areas such as the zona incerta, cerebral peduncle (crus cerebri), lenticular fasciculus, and the medial lemniscus are also included in this area. The optic tract is included in this area in the most anterior extent.”* – Neuromorphometrics (8)

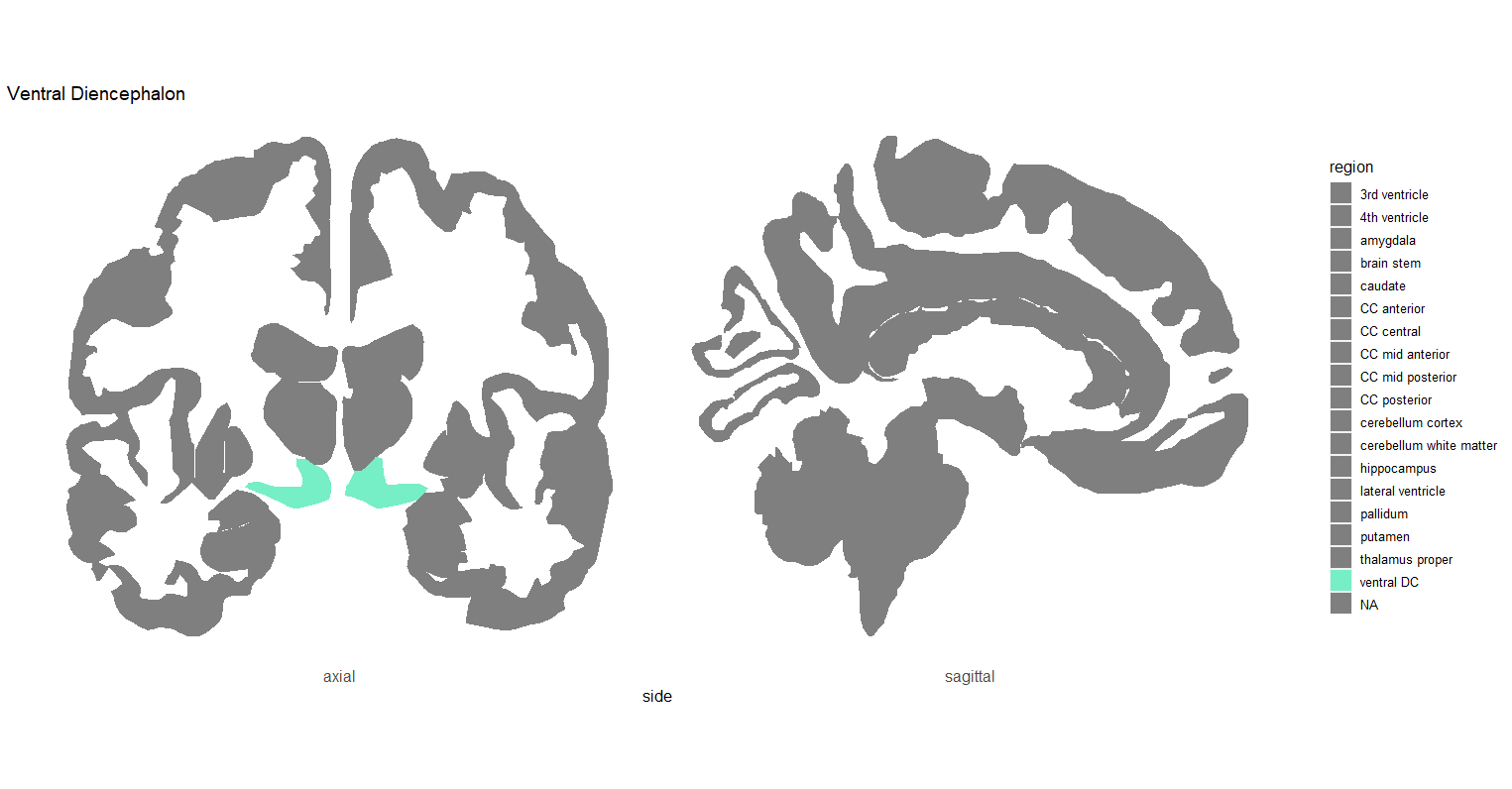

**Supplemental Figure 1**: Map demonstrating the position of the FreeSurfer region the Ventral Diencephalon (in green), in a representative coronal section of the human brain.

MRI scanning in UK Biobank – further information:

The FreeSurfer region the ‘temporal pole’ was not included in the UKB parcellation data, so was not included in the UKB analysis or the mega-analysis.

Supplemental Information References:

1.	Schmidt MR, Narayan AJ, Atzl VM, Rivera LM, Lieberman AF. Childhood Maltreatment on the Adverse Childhood Experiences (ACEs) Scale versus the Childhood Trauma Questionnaire (CTQ) in a Perinatal Sample. Journal of Aggression, Maltreatment & Trauma. 2020;29(1):38-56.

2.	Paivio SC, Cramer KM. Factor structure and reliability of the Childhood Trauma Questionnaire in a Canadian undergraduate student sample. Child Abuse & Neglect. 2004;28(8):889-904.

3.	FreeSurfer. FreeSurfer Wiki: Subcortical Segmentation [Available from: <https://surfer.nmr.mgh.harvard.edu/fswiki/SubcorticalSegmentation>.

4.	Filipek PA, Richelme C, Kennedy DN, Caviness VS, Jr. The young adult human brain: an MRI-based morphometric analysis. Cereb Cortex. 1994;4(4):344-60.

5.	Makris N, Oscar-Berman M, Jaffin SK, Hodge SM, Kennedy DN, Caviness VS, et al. Decreased Volume of the Brain Reward System in Alcoholism. Biological Psychiatry. 2008;64(3):192-202.

6.	Seidman LJ, Faraone SV, Goldstein JM, Goodman JM, Kremen WS, Matsuda G, et al. Reduced subcortical brain volumes in nonpsychotic siblings of schizophrenic patients: a pilot magnetic resonance imaging study. American Journal of Medical Genetics. 1997;74(5):507-14.

7.	Fischl B, Salat DH, Busa E, Albert M, Dieterich M, Haselgrove C, et al. Whole brain segmentation: automated labeling of neuroanatomical structures in the human brain. Neuron. 2002;33(3):341-55.

8.	Neuromorphometrics. Neuromorphometrics Segmentation: Ventral Diencephalon [Available from: <http://neuromorphometrics.org:8080/seg/html/segmentation/ventral%20diencephalon.html>.
